# Psychological impact of COVID-19 on frontline healthcare workers during the early months of the pandemic and responses to reduce the burden, helping to prepare for Disease X: A systematic review

**DOI:** 10.1101/2023.11.28.23299078

**Authors:** Jarryd S. Ludski

## Abstract

**Objective:** The COVID-19 pandemic placed enormous strain on healthcare workers (HCW) and systems. With currently over 766 million cases, a high risk of workplace-acquired infection and a constantly evolving disease trajectory, COVID-19 placed an incredible burden on frontline HCWs. Studies from previous pandemics highlight significant psychological distress in these workers, yet mental health remained a secondary consideration in many hospitals pandemic response. This review explores the psychological impact of COVID-19 on frontline HCWs during the early stages of the pandemic and describes responses implemented by health services to reduce this impact. Additionally, it aims to provide a framework for future evidence-based programs that support the wellbeing of frontline HCWs throughout the ongoing pandemic and into the future, helping to prepare for Disease X.

**Methods:** A systematic review was completed using MEDLINE, CINHAL and Cochrane databases with bibliographic and grey literature searches.

**Results:** 17 publications were included. Symptoms of psychological distress were reported in up to 70% of frontline HCWs, with as many as 50% suffering depression, 62% reporting anxiety and 45% of those requiring quarantine experiencing insomnia. Mindfulness training, safe rest areas, mental health practitioners and pandemic rostering are responses that have been implemented across health services during the pandemic, but their efficacy in reducing psychological burden has not been fully assessed.

**Conclusions:** The impact of COVID-19 has been enormous; however, its final toll remains unknown. High rates of psychological distress amongst frontline HCWs means the impact will extend far beyond the virus itself. Health services must implement evidence-based resilience strategies to ensure the safety of their frontline staff now and into the future.

## Introduction

In late 2019, the novel coronavirus (COVID-19) was first recognised and by the 30^th^ of January 2020, it was declared a public health emergency of international concern^1^. Ten months later, on the 27^th^ of October 2020, global case numbers exceed 43 million, with over 1.1 million deaths^2^ including many healthcare workers (HCW). By May 2023, there have been over 766 million cases globally, with almost 7 million deaths^3^. The threat of COVID-19 posed a great challenge to hospital systems, leading to staff shortages, scarcity of medical supplies and an increased risk of workplace acquired infections^4^. In September 2020, the World Health Organisation (WHO) announced that despite healthcare workers accounting for only 3% of the population, they have accounted for 14% of COVID-19 cases in many countries and as high as 35%^5^. COVID-19 was a worldwide health crisis, yet it is not the first pandemic faced by the modern world. Over the last century, the Spanish Flu, Severe Acute Respiratory Syndrome (SARS), Middle East Respiratory Syndrome (MERS), Ebola and Swine Flu have all emerged^6^, nor will it likely be the last. The WHO identifies ‘Disease X’ as a priority for global investment, research and development indicating a currently unknown pathogen with the potential to cause a serious international epidemic^7^ Literature from these previous pandemics highlight that apart from the physical morbidity and mortality, these events cause significant mental health burden including anxiety, depression, burnout, insomnia, and post-traumatic stress disorder (PTSD) amongst frontline HCWs^6^. Despite this knowledge and previous experience, the mental health of frontline HCWs often remains a secondary consideration behind the needs of the public. The psychological, emotional, and physical demands placed on overstretched frontline workers may cause long-lasting and substantial negative impacts on wellbeing^8^ that stretch far beyond COVID-19.

### Review Objectives

The primary aims of this study are to:

a. Review and describe the available literature on the psychological symptoms, burden, and impact of the early stages of the COVID-19 pandemic on frontline HCWs.
b. Highlight responses implemented by health services to mitigate the psychological symptoms, burden, and impact on frontline HCWs.
c. To provide a framework for healthcare services to design and initiate evidence-based programs to support the wellbeing of their frontline workers throughout a pandemic
d. Provide a framework for early initiation of programs to reduce similar burden faced with a potential Disease X.

## Methods

### Study Design

This systematic review was designed following discussions with senior emergency department physicians, nurses, and administration staff, with a broad research question developed to attempt to gain a greater understanding of the broad impact of COVID-19 on frontline HCWs.

### Ethics Approval

Ethical approval was deemed not required, as all data was retrieved from previously published studies, where approval was obtained by primary investigators.

### Search Strategy

Published studies were found by search MEDLINE (OVID interface), Cumulative Index to Nursing and Allied Health Literature (CINAHL) and Cochrane Central Register of Controlled Trials (Cochrane Library). Additional literature was identified through reference tracking and grey literature searches. Additional bibliographic and grey literature search of the official WHO website was also conducted. Databases were searched for the following terms and included articles up to October 2020:

**SEARCH STRATEGY**

‘impact’

**AND**

‘Covid-19’ OR ‘coronavirus’ OR ‘2019-ncov’ or ‘sars-cov-2’ or ‘cov-19’

**AND**

‘Emergency department’ OR ‘emergency room’ OR ‘accident and emergency’ OR ‘accident & emergency’ OR ‘a&e’ OR ‘a & e’

**AND**

‘staff’ OR ‘healthcare workers’ OR ‘nurses’ OR ‘medical workers’ OR ‘medical practitioners’ OR ‘medical workforce’ OR ‘healthcare professionals’.

### Types of Studies

Primary empirical research studies will be eligible for inclusion, while editorials, protocols for planned studies, abstracts and dissertations were excluded.

### Inclusion criteria

All relevant articles published or translated in English, with full text available will be included for review.

### Screening procedure

All three-step screening process of the title, abstract, and full-text review was undertaken by the author. First screening of titles and abstracts for relevance was completed. Secondly full texts were obtained for all potentially relevant publications and reviewed. Finally, all relevant publications were selected for inclusion in this review. This process is documented as a preferred reporting item for systematic reviews and meta-analysis (PRISMA) flowchart below (Figure 1).

**Figure 1:**
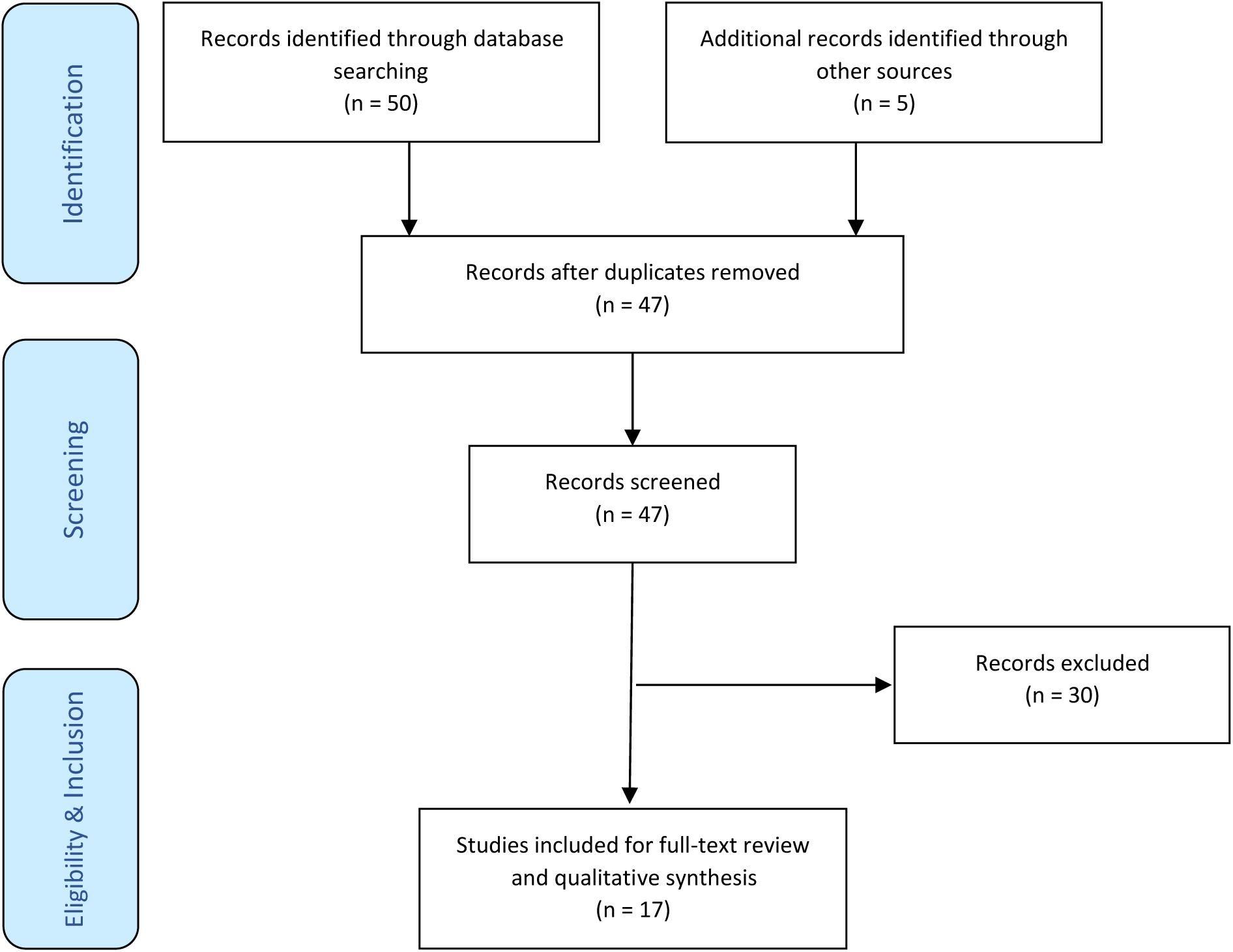
Search Strategy flow diagram

### Data extraction

The included studies were charted into a customized data extraction form to extract all relevant data from each study. Data extraction was performed by the author (JL), with the following data extracted.

- First author
- Year of publication
- Year of data collection
- Study design
- Country of data collection
- Study population and size of population
- Cohort occupation
- Outcome measures used (e.g. questionnaires, online survey, etc.)
- Rates of psychological, physical, and emotional symptoms
- Impact on frontline HCWs

## Results

A total of 55 publications were collected. Following removal of duplicates, 47 publications underwent abstract review with 17 publications deemed relevant for inclusion in our study as outlined in **figure 1**.

### Mental Health impacts of COVID-19 reported in Frontline HCW

COVID-19 has posed a huge threat to public health and placed frontline HCWs at significant risk of psychological distress, with the literature highlighting the large proportion suffering ill-effects of mental disorders. Up to 70% of HCWs^6,11^ have reported symptoms of anxiety, depression, PTSD, sleep disturbances, cognitive decline, or burnout.

#### Depression

Recent studies undertaken during the COVID-19 pandemic have found a high prevalence of depressive symptoms in frontline HCWs. Depression was reported in 44% of 1103 frontline nurses^9^, with 35% reporting moderate or severe symptoms. Furthermore, a study of 4,679 doctors and nurses across 348 Chinese hospitals highlighted a 35% prevalence of depressive symptoms^10^.

Another study reported symptoms in 44% of HCWs including physicians, medical residents, nurses, technicians, and public health professionals^11^. Furthermore, 37% of HCWs in quarantine or working directly with COVID-19 patients report suffering from depressive symptoms^12^. Finally, some COVID-19 studies have reported depression in as high as 50% of HCWs, showing the impact to be greater than that found during the SARS pandemic^6^.

#### Anxiety

Global studies have reported symptoms of anxiety in 44-62% of frontline HCWs, with a quarter suffering moderate to severe anxiety^6,11^. A study in India highlighted that 60% of HCWs in quarantine or working with COVID-19 patients experienced anxiety symptoms^12^. Moreover, a large proportion of respondents reported anticipatory anxiety^13^, with HCWs holding concerns regarding the future given the disease trajectory and virus transmission. Additionally, 75-80% of HCWs have felt psychologically unsafe and held concerns about being a source of COVID-19 infection for their families as well as failing to provide their usual standard of care to colleagues or friends^14,15^.

#### PTSD

While the risk of PTSD in frontline HCW has long been established under standard working conditions, it is even greater during times of natural disaster such as in the COVID-19 pandemic where the stress and trauma are repeated daily^10^. Studies from the SARS pandemic demonstrated symptomatic PTSD in up to 33% of nurses who worked in intensive care units^6^, and greater symptomatology in those who experienced quarantine or worked in high-risk units compared with their colleagues in low-risk units^6,11,16^. Finally, the response to the current pandemic demonstrates many characteristics of mass trauma, with individuals moving into a state of hypervigilance, manifesting avoidance strategies, experiencing negative moods and culminating in psychological distress^12^.

#### Insomnia

There is currently limited evidence available on insomnia in HCWs, however it has been reported in 29% of frontline workers and in up to 45% of those required to quarantine or working directly with confirmed positive cases^6,11,12^.

#### Burden of Personal Protective Equipment

Throughout the world, the mandatory use of personal protective equipment (PPE) has been a mainstay in the fight to control COVID-19 infections in HCWs. For HCWs this involves donning a close-fitting N95 face mask, protective eyewear, a gown, and surgical gloves. It is often cumbersome, uncomfortable and makes drinking water and breathing difficult^17^. This, in combination with fear over limited supply of PPE and confusion about its effectiveness, heightens work intensity and increases feelings of fatigue that have detrimental effects on mental health^4,6^. Moreover, the use of PPE has been associated with headaches that decrease work performance and productivity when used for greater than 4 hours per day^17^.

#### Measures implemented to assist frontline healthcare workers and reduce the burden

As outlined, frontline HCWs are at a considerable risk of psychological distress, throughout the Covid-19 pandemic and beyond. The psychological burden can lead to significant detrimental effects on their work, such as poor morale, staff conflicts, absenteeism, apathy and lapses in care^9^, which increase patient dissatisfaction and may result in poor patient outcomes. Staff wellbeing is an integral pillar in a hospital’s pandemic response^15^ with multiple strategies being suggested and implemented throughout the world in response to the pandemic.

#### Psychiatric support/Support by Mental Health Practitioners

Healthcare services have utilised consultations with mental health experts, distribution of health-promoting webinars and stress management training as a means of reducing the mental health burden faced by frontline HCWs^6^. With the majority of HCWs reporting mild symptoms of psychological distress, interventions such as activity scheduling, behavioural activation, and relaxation techniques may be beneficial^12^. Throughout Italy, access to a hotline with psychiatric support has been offered in major hospitals, along with the implementation of acceptance and commitment therapy (ACT). ACT is a mindfulness-based cognitive behavioural therapy aimed at improving psychological flexibility and has been shown to be effective in patients with PTSD^18^. An ACT-based psychoeducation booklet has also been designed for HCWs^10^. Finally, peer supporters trained in psychological first aid have been used in emergency departments to promote natural recovery from crisis and traumatic events, along with wellbeing drop-in sessions providing additional avenues for vulnerable staff to seek support^15^.

#### Leadership and Effective communication

Anticipatory anxiety, fear, misinformation, and a lack of confidence in PPE have been linked to significant psychological distress in frontline HCWs^6^. However, with clear, identifiable leadership and effective communication, anxiety can be minimised^15^. Moreover, through ensuring regular updates, opportunities to ask questions and extending support beyond just hospital issues to areas such as family needs or finances, feelings of helplessness experienced by HCWs can be reduced, alleviating stress, and improving mental wellbeing^6,15^.

#### Rostering

COVID-19 posed a unique set of challenges to rostering in emergency departments. To ensure they remained staffed with appropriately skilled clinicians despite the possibility of staff being furloughed or isolating, many hospitals adapted their rosters. The pandemic strategy in Singapore’s largest tertiary teaching hospital involved dividing staff into five equally balanced teams. They worked 12-hour shifts with handovers and overlapping staff kept as brief as possible^19^. The longer shifts built-in buffer capacity that provided additional rest days if no teams required quarantining and meant that the average hours worked would only slightly increase if as many as three teams were required to isolate^19^. In a large Melbourne metropolitan hospital, a roster was also implemented where full-time staff worked no more than four consecutive shifts followed by three days off, with staff rotating between areas of high and low stress^15^. This ensured optimal recovery time for staff, protecting against chronic stress and maintaining staff capacity to fulfil their roles^15^. Moreover, four-hourly breaks were taken as encouraged by the Australian College of Emergency Medicine (ACEM) and the final 30 minutes of shifts protected for debriefing and reflective self-care^15^. Finally, some hospitals redeployed nurse practitioners and physician assistants to areas of critical need, providing support with low acuity diagnoses, discharging patients and collaborating with telehealth physicians to reduce the burden on frontline workers^20^.

#### Safe rest areas

Research from China indicates the need for a ‘COVID-safe’ rest area for HCWs working in high-risk areas^10^. At Royal Melbourne Hospital, the Emergency Department has expanded non-clinical areas to provide these safe places for rest, mindfulness, yoga and sustaining social connectedness^15^.

Additionally, safe areas provide easy access to water, and educational material for HCWs to support psychological wellbeing^10^.

## Discussion

This study has highlighted the enormous impact of the early stages of the COVID-19 pandemic on the psychological health of frontline HCWs globally. Over 50% are reporting symptoms of psychological distress such as anxiety, depression, insomnia, burnout, or acute stress reactions^11^. Consequently, this pandemic has the potential to derail career paths, decrease job satisfaction, accelerate compassion fatigue, and cause significant detriment to patient outcomes. Additionally, those who work in emergency departments, directly with COVID-19 patients, or are forced to quarantine, are at a significantly higher risk of distress, making the volume of affected HCWs potentially huge^4,6,9,11,21^; consequently, reducing a healthcare service’s ability to function and placing their non-COVID patients at risk. Furthermore, psychological distress has been linked to medical errors, delayed recoveries, and poor patient satisfaction. Therefore, the implementation of evidence-based strategies to reduce psychological distress is crucial to ensure the health of frontline workers and improve patient care. Additionally, these interventions will improve work satisfaction and productivity, reduce absenteeism and employee turnover, and assist in the formation and maintenance of a supportive, safe, and effective work culture^22^.

The establishment of evidence-based, preventive, and supportive measures to improve the mental health of frontline HCWs should be a priority for all healthcare services in the early stages of a pandemic. Many have already met this challenge by putting in place measures they believe will reduce the burden. While this is a positive step for HCWs, unfortunately, the effectiveness of these programs has only been demonstrated during non-pandemic times and has not yet been rigorously explored during a pandemic.

Resilience is defined as the maintenance or quick recovery of mental health during or after periods of stressful exposure^23^. A Cochrane review into resilience training in healthcare professionals, has highlighted a reduction in symptoms of depression and levels of stress when based on the principles of mindfulness, cognitive-behavioural therapy, and ACT^23^. Additionally, yoga-based interventions have been shown to reduce emotional exhaustion, burnout and anxiety whilst improving sleep and mental health^22^. Moreover, mindfulness practices such as weekly 30-minute sessions, structured programs, and the use of apps such as Smiling Mind, have further demonstrated reductions in perceived stress, anxiety levels and depressive symptom^22,24,26,27^. These strategies have been recommended by ACEM and the UK Intensive Care Society to improve staff wellbeing during and beyond COVID-19^28,29^.

Other strategies initiated such as the use of mental health professionals, peer-support groups and ensuring effective, open communication, have limited evidence in the literature^30^. However, a supportive supervisor and work culture, along with practical support have been shown to protect staff mental health^31^, Despite the lack of evidence, both the ACEM and ICS recommend these strategies, when endorsed by strong leadership, as feasible and sustainable measures to support staff wellbeing^26,29^.

The implementation of policies and practices to improve the psychological health of HCWs, have been shown to be positively associated with increased job satisfaction, improved workplace culture and better patient outcomes^32^. Patient mortality rates, readmission rates, adverse events and patient satisfaction are all improved with a positive workplace culture^32^. The adoption of health-promoting strategies is therefore a crucial tool in the arsenal of any health service to support their staff and patients throughout the pandemic and into the future.

### Future Directions

Future work should focus on ensuring all health services are aware of the high risks facing their frontline HCWs and evidence-based interventions that can be implemented to support psychological wellbeing. This could be achieved through the distribution of informative flyers, such as the example shown in appendix 1. Further research is indicated to investigate the long-term prevalence of psychological distress in individual hospitals and its associated burden. With the large variation in the amount of COVID-19 exposure faced by each hospital, the true burden should be identified, along with the quantitative evaluation of measures implemented to improve mental health and resilience. This could be achieved through the confidential distribution of anonymous pre- and post-intervention questionnaires that address symptoms of psychological distress (appendix 2).

### Limitations

The study was limited by heterogeneity in global study populations and variations in symptom evaluation tools.

## Conclusion

The impact of the COVID-19 pandemic is enormous, with its true toll potentially always unknown. So far, over 766 million cases have been recorded worldwide with almost 7 million deaths. While the spread is now more contained, frontline HCWs are charged with continuing to ensuring the health needs of this patient population are met, recovering, and preparing for the next pandemic, disease X. The psychological burden placed on those frontline workers during the early stages of the pandemic is another crisis that must be addressed. With over 50% of HCWs reporting symptoms of psychological distress, the impact of the pandemic will extend far beyond the virus itself. Health institutions have implemented a multitude of strategies to assist in minimising the burden placed on frontline workers. These resilience strategies may be costly, and their use should be rigorously tested for efficacy as we move into the unknown post-COVID and COVID-normal world. The use of validated strategies will provide a crucial tool in supporting the mental health of frontline staff, allowing them to continue their much-needed work and ensuring the safety of their entire communities now and into the future.

## Data Availability

All data produced in the present study are available upon reasonable request to the authors

## Author Contributions

Jarryd Ludski: Project proposal, conducted literature search and systematic review, drafted manuscript and revisions, final approval of the manuscript for submission.

## Acknowledgments

I would like to acknowledge Oliver Hoffman, who assisted in designing our informative flyer for distribution to health services and the hard work and dedication shown by frontline HCWs throughout the COVID-19 pandemic.

## Guarantor

Dr. Jarryd Ludski

---

**Appendix 1:**
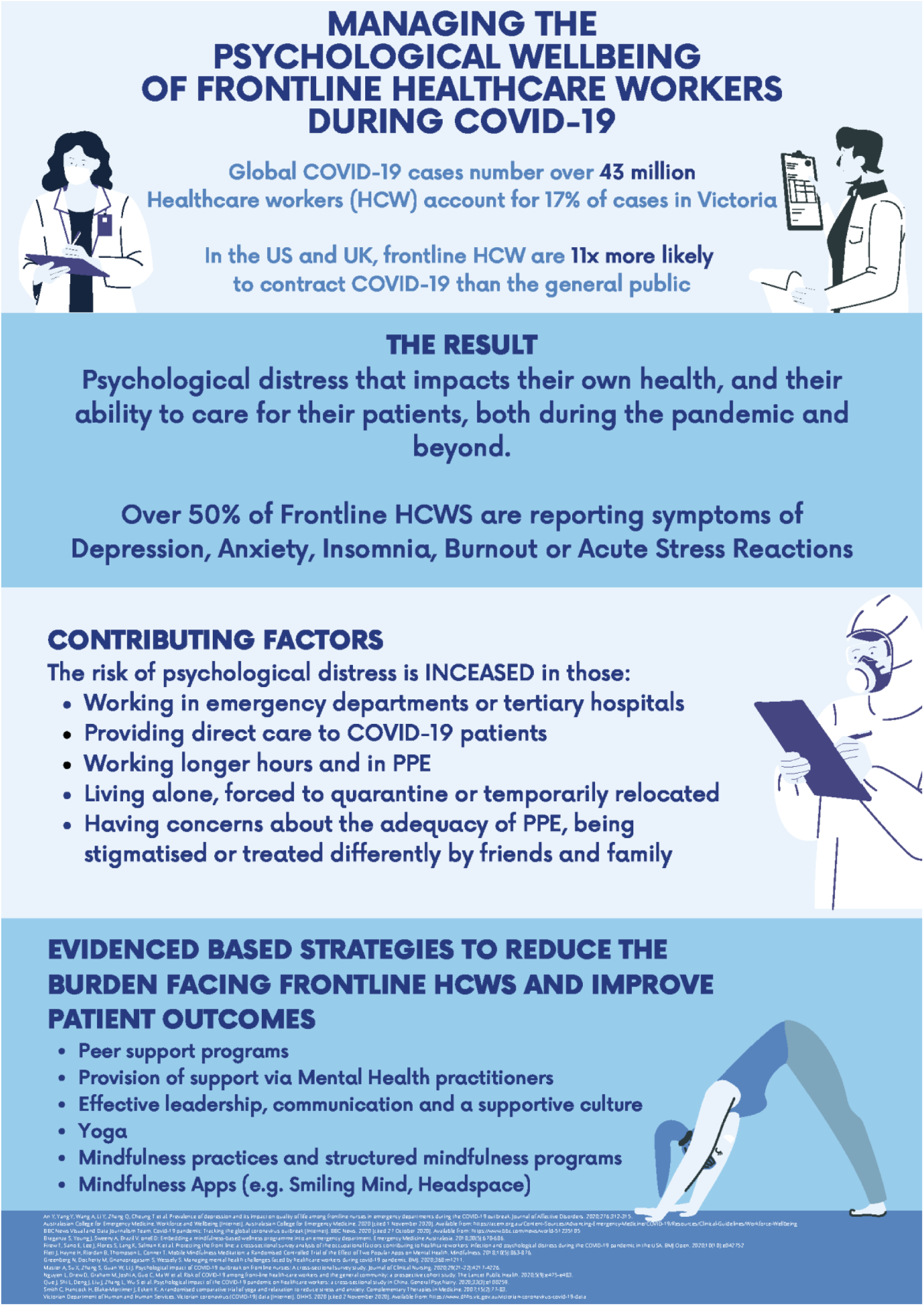
Example of an informative flyer to distribute to and amongst healthcare services, using COVID-19 as an example.

**Appendix 2:**
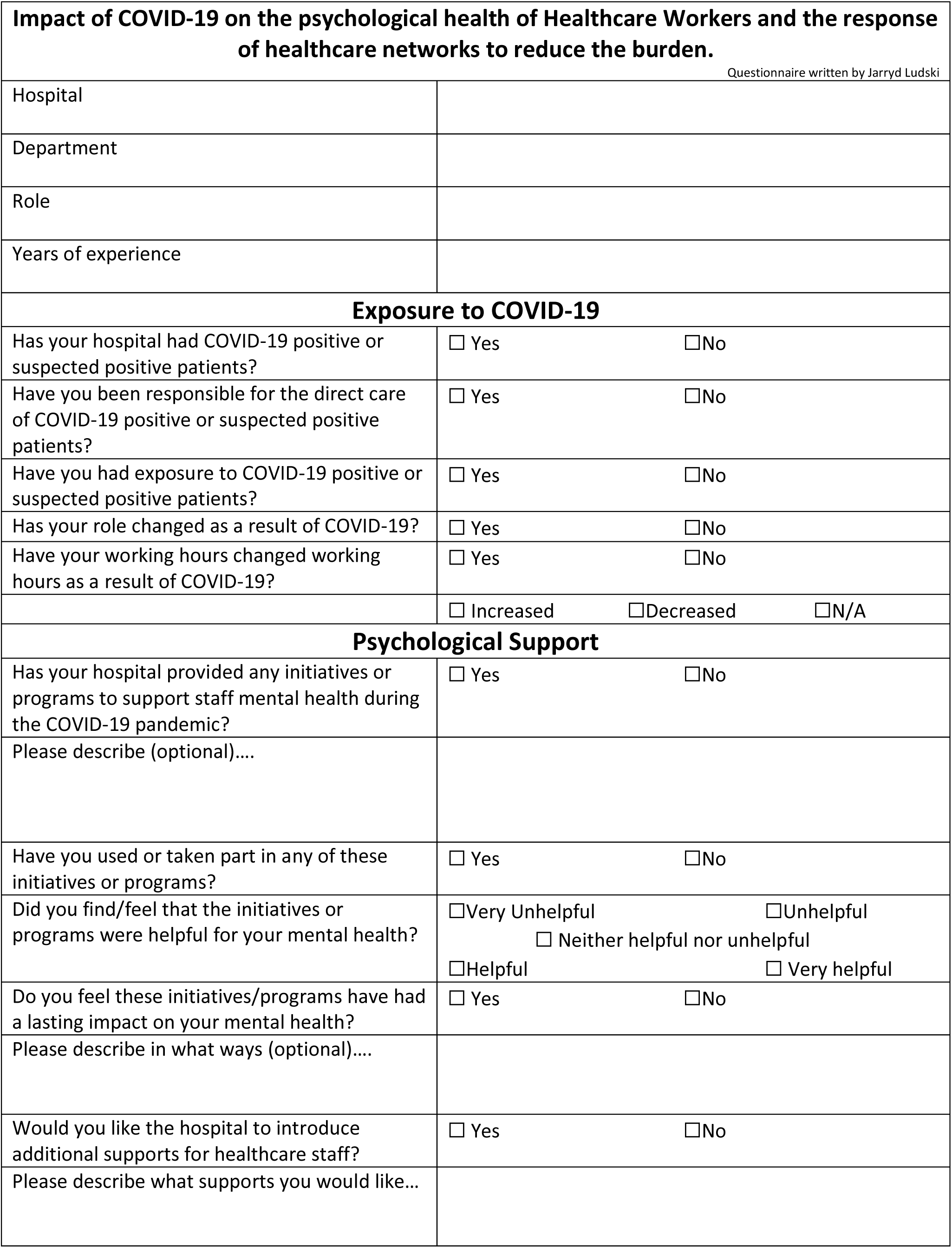
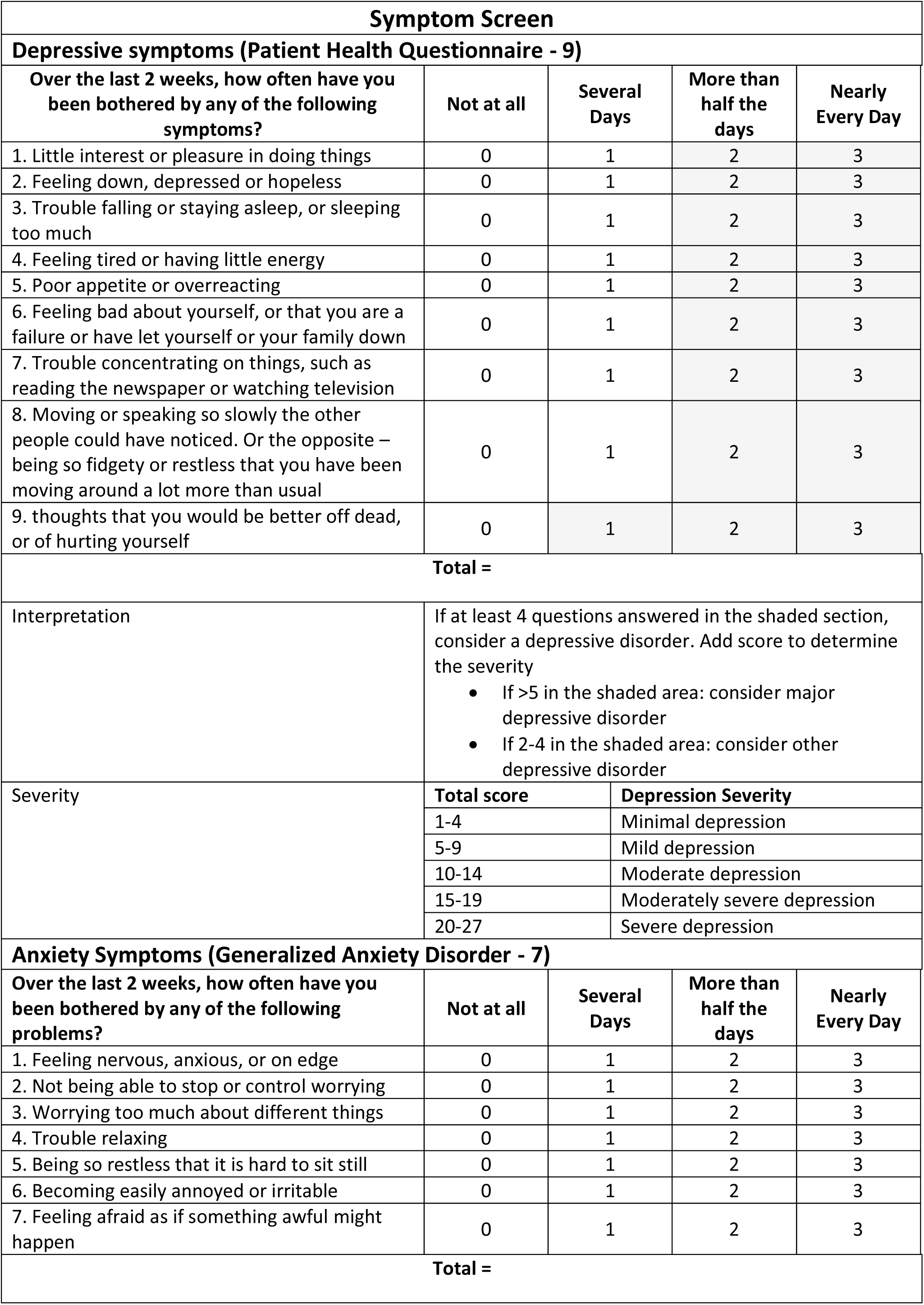
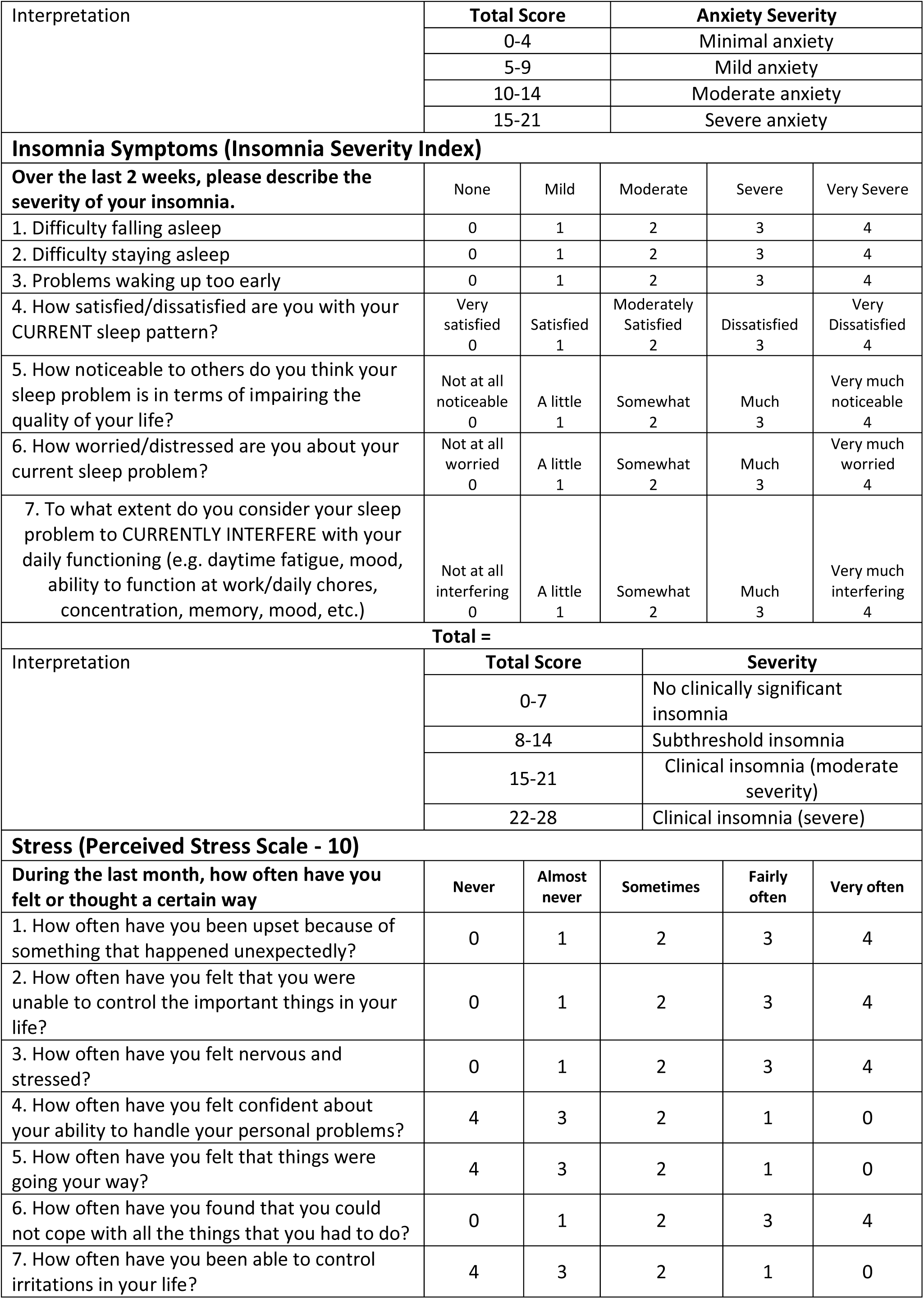
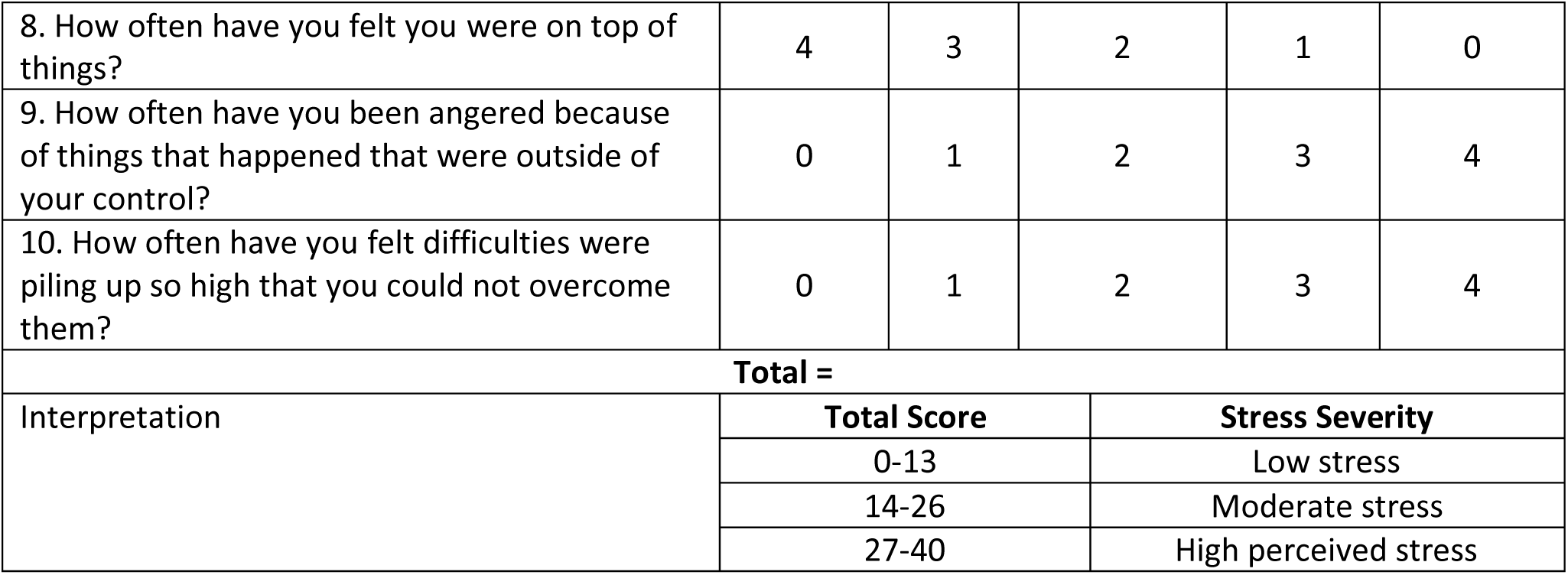
Example questionnaire for evaluation of the baseline psychological health of frontline HCWs and subsequent effectiveness of implemented interventions, using COVID-19 as an example.

